# Robustness of plasma p-Tau217 diagnostic thresholds for Alzheimer’s disease across various clinical populations and laboratory environments

**DOI:** 10.64898/2025.12.19.25342412

**Authors:** Jean-Louis Bayart, Nicolas Villain, Vincent Planche, Emilien Boyer, Lise Colmant, Isabelle Le Ber, Gaëtane Picard, Fabienne Clot, Stéphanie Bombois, Hassan El-Mazria, Giulia Dingeo, Sarra Bahroun, Fatimah Nabeebaccus, Lara Huyghe, Thomas Gerard, Lisa Quenon, Yasmine Salman, Emmanuelle Durand, Aurélie Bedel, Sophie Auriacombe, Renaud Lhommel, Clara David, Pascal Kienlen-Campard, Adrian Ivanoiu, Jonathan Douxfils, Richard Levy, Foudil Lamari, Bernard J. Hanseeuw

## Abstract

**Background and Objectives:** Blood-based biomarkers (BBMs), especially tau phosphorylated at threonine 217 (p-tau217), offer a minimally invasive approach to diagnose Alzheimer’s disease (AD) with strong potential for clinical implementation. However, the robustness and transferability of diagnostic thresholds across sites remain uncertain. This study aimed to evaluate BBMs’ ability to predict amyloid and tau status in a deeply phenotyped AD research cohort and to test p-tau217 cutoffs across diverse clinical populations and analytical instruments.

**Methods:** We included four Western European cohorts (total n=411): an exploration cohort from Brussels with amyloid and tau-positron emission tomography (PET) data (n=215), and clinical validation cohorts from Paris (memory clinic, n=117, and monogenic FTLD, n=43) and Bordeaux (early-onset neurodegenerative disorders, n=36). BBMs concentrations (p-tau217, p-tau181, NfL, BD-tau, Aβ42, Aβ40) were measured. Of these, p-tau217 was measured at each site using three Lumipulse analyzers (G600II in Brussels and Bordeaux; G1200 in Paris). Amyloid status was determined by PET or CSF Aβ42, and tau status by tau-PET or CSF p-tau181.

**Results:** Plasma p-tau217 outperformed other BBMs, and their related ratios. In the exploration cohort, p-tau217 closely related to amyloid and tau PET, clearly separating Aβ+ from Aβ− individuals (AUC=0.96; optimal cutoff 0.193 pg/mL with ∼92% sensitivity/specificity) and tracking tau-PET Braak stages (AUC=0.93–0.98). Using amyloid-derived thresholds, p-tau217 detected medial temporal and neocortical tauopathy with >90% sensitivity and specificity at stricter dual 95–97.5% sensitivity/specificity cutpoints (0.142/0.256 pg/mL and 0.110/0.319 pg/mL). In validation cohorts, the 0.193 pg/mL cutoff accurately discriminated clinical-biological AD from non-AD in routine care (AUC=0.98; ∼93% sensitivity/specificity), in early-onset dementia (AUC=0.94; >92% sensitivity/specificity), and in the monogenic FTLD cohort (89% specificity). In routine care, gray zones (8–25%) were largely resolved with second-line CSF testing (>75%). Elevated p-tau217 was rare in non-AD, mainly in FTLD-ALS and some older individuals with possible AD copathology. p-tau217/Aβ42 and p-tau217/Aβ42/Aβ40 ratios added no diagnostic value.

**Discussion:** p-tau217 thresholds maintained high diagnostic accuracy (>90%) across independent sites, analytical platforms, and various clinical situations, supporting their robustness and transferability. These observations support implementing p-tau217 as a reliable, scalable test for detecting AD pathology, for diagnostic work-ups of patients with objective cognitive impairment, not for general population screening.

## INTRODUCTION

Alzheimer’s disease (AD) is a neurodegenerative disorder characterized by progressive cognitive decline leading to loss of autonomy, and accumulation of β-amyloid and tau aggregates in the brain parenchyma at autopsy, along with synaptic and neuronal loss.^1^ Blood-based biomarkers (BBMs) are reshaping AD diagnostics by providing scalable, minimally invasive, and cost-effective tools to detect and monitor underlying pathology in both research and clinical practice.^2,3^ Among these, the plasma concentration of phosphorylated tau at threonine 217 (p-tau217) has emerged as a leading BBM for β-amyloidosis and tau pathology, with diagnostic accuracy comparable to CSF and PET and superior to other plasma measures, including p-tau181.^4–7^ Additional BBMs such as the Aβ42/Aβ40 ratio, neurofilament-light chain (NfL) and Brain-Derived Tau (BD-Tau) capture complementary yet distinct aspects of AD pathophysiology, from amyloid aggregation to neuronal injury and neurodegeneration.^8–12^

Clinical implementation of plasma p-tau217 is challenged by assay performance, interplatform variability, pre-analytical conditions, and cutoff definitions. On the Lumipulse^®^ chemiluminescent enzyme immunoassay platform (Fujirebio, Gent, Belgium), 95% sensitivity/specificity (Se/Sp) cutoffs to highlight Aβ status range from 0.124–0.328pg/mL to 0.252–0.443pg/mL, reflecting differences in study populations, reference standards, and analytical procedures.^13–15^ This heterogeneity raises key questions regarding cross-site applicability of thresholds and the need for locally derived decision limits.

It is also debated whether a single biomarker approach is sufficient or whether composite biomarkers, i.e. particularly the p-tau217/Aβ42 ratio; are required to improve robustness and reduce indeterminate “gray zone” classifications.^16^ Emerging studies suggest that such ratios may improve diagnostic precision or reduce gray zones in heterogeneous or comorbid populations, possibly by mitigating p-tau217-related variability and systemic confounders.^10,17^

To support broad clinical implementation of plasma p-tau217, three conditions are crucial: (i) demonstration that p-tau217 thresholds can be transferred across independent analyzers and laboratories; (ii) confirmation that these thresholds maintain high performance in various clinical populations; and (iii) integration of p-tau217 into simple two-step diagnostic pathways specifying when CSF analyses or PET imaging remain necessary. This is particularly important for centers lacking resources for extensive local validation, which requires robust externally validated thresholds to guide diagnostic decisions and facilitate access to disease-modifying therapies.

In the present study, we first compared the clinical performance of p-tau217, p-tau181, Aβ42/Aβ40, NfL, BD-Tau and their derived ratios for predicting brain β-amyloidosis in a deeply phenotyped research cohort of 215 individuals with amyloid and tau-PET data. We then selected the best-performing marker, p-tau217, and tested the robustness and transferability of amyloid-anchored Lumipulse-based thresholds across three independent validation cohorts: (i) a routine memory clinic cohort from Paris; (ii) a cohort of patients with early-onset neurodegenerative dementia from Bordeaux and (iii) a monogenic frontotemporal lobar degeneration (FTLD) cohort from Paris. We assessed amyloid-anchored p-tau217 thresholds’ ability to detect tau-PET positivity (including regional patterns in the medial temporal lobe and neocortex) and to classify clinical-biological AD versus non-AD diagnoses. We finally examined a two-step algorithm combining plasma p-tau217 with second-line CSF/PET to resolve gray-zone cases.

## METHODS

### Participants

#### Exploration research cohort (Brussels, Belgium)

Clinically unimpaired (CU) individuals, enriched for apolipoprotein E (APOE) ε4 carriers, were recruited through local advertising. Patients with mild cognitive impairment (MCI) and dementia (DEM) were recruited from the Memory Clinic of Cliniques Universitaires Saint-Luc (the academic hospital of UCLouvain, Brussels, Belgium) between November 1, 2020 and November 30, 2024 (Eudra-CT number: 2018-003473-94). Each participant underwent a comprehensive neuropsychological assessment (MMSE, episodic memory, language, visuospatial, and executive functions). MCI was defined as MMSE ≥ 24/30 and cognitive impairment (z-score <−1.5) in at least one cognitive domain (language scores disregarded if testing was not conducted in the native language of the participant) without significant functional impairment.^18^ Dementia diagnosis required MMSE score < 24 and functional impairment. For statistical analyses, MCI and dementia cases were pooled together as cognitively impaired (CI).

#### Validation cohort: clinical routine care (Paris, France)

The Paris prospective cohort included consecutive patients referred to the Institute of Memory and Alzheimer’s Disease (IM2A), Pitié-Salpêtrière Hospital, Assistance Publique–Hôpitaux de Paris (AP-HP), for evaluation of cognitive or behavioral complaints between January 29, 2024, and July 4, 2024, for whom CSF sampling was clinically indicated. All patients underwent neurological and neuropsychological assessment, structural neuroimaging, and routine blood tests to screen for reversible causes of cognitive impairment. Lumbar puncture for CSF AD biomarkers analysis was performed, strictly within routine clinical care. Residual EDTA plasma and CSF samples were analyzed blindly at the Pitié-Salpêtrière Biochemistry

Department. Patients were categorized based on locally validated CSF Aβ42 and p-tau181 cutoffs into amyloid and tau positivity (A/T) profiles. Patients were classified as having AD if diagnosed with probable or highly probable clinical-biological AD per IWG 2021 criteria.^19^ The attending physician could also identify comorbid pathologies likely contributing to the phenotype. For this study, clinical-biological AD was defined as any AD diagnosis, primary or comorbid. Non-AD corresponded to other neurodegenerative, mixed, or non-neurodegenerative etiologies.

#### Validation cohort: early onset neurodegenerative dementia (Bordeaux)

BioNEODEM, an ancillary study of the NEODEM cohort (NCT04254094), enrolled individuals with early-onset neurodegenerative dementia at Bordeaux University Hospital. Inclusion criteria were: new diagnosis of a neurodegenerative disease within 6 months; onset of first relevant symptoms before age 65; and CDR 0.5–1 at baseline. Clinical diagnoses included early-onset AD (EOAD), behavioral variant frontotemporal dementia (bvFTD), primary progressive aphasia (PPA) variants, progressive supranuclear palsy (PSP) and corticobasal degeneration (CBD). Genetic testing was performed at the discretion of the physicians and the participants or their families: among 36 patients, 6 patients with EOAD and 6 patients with FTD underwent genetic testing for an autosomal dominant cause, and only one GRN mutation was identified in a bvFTD patient.

#### Monogenic FTLD cohort (Paris)

The monogenic frontotemporal lobar degeneration (FTLD) cohort included patients investigated at the IM2A and collaborating centers, referred for genetic testing due to FTLD-spectrum phenotypes. Surplus diagnostic blood samples were centralized. Only individuals with pathogenic or likely pathogenic variants in *C9ORF72*, *GRN*, or *MAPT* were included. Clinical characterization depth varied, but all fulfilled accepted FTLD-spectrum criteria; FTLD–ALS overlap presentations were captured.

##### Determination of Aβ (A) status

In Brussels, amyloid-PET with [^18^F]flutemetamol or [¹¹C]PIB followed standard protocols; scans >25 Centiloids were considered Aβ+.^20,21^ When PET was unavailable, CSF Aβ42 measured on Lumipulse G600II defined Aβ status (Aβ+ if ≤544 pg/mL).^22^ In Paris and Bordeaux, CSF Aβ42 was measured on Lumipulse G1200 and G600II analyzers respectively, and locally validated cutoffs were applied (<650 pg/mL in Paris; 600 pg/mL in Bordeaux).^23^ When the CSF Aβ42 and p-tau181 results were discordant (A-/T+ or A+/T-), the Aβ42/40 ratio was also measured to confirm the A status.

### Determination of Tau (T) status

In Brussels, tau status was established using [^18^F]MK-6240 Tau-PET. [18F]-MK-6240 Tau-PET images were co-registered with a 3D-T1 MRI using the PetSurfer pipeline, a set of tools within FreeSurfer for end-to-end integrated MRI-PET analysis. Standardized Uptake Value ratio (SUVr) values were extracted for all regions from the Desikan-Killiany Atlas using cerebellum gray matter as a reference region. A meta-temporal region of interest (ROI) that included the amygdala, the hippocampus, the entorhinal, parahippocampal, fusiform, inferior temporal, and middle temporal cortices was then calculated.^24–26^ Two experienced nuclear physicians (T.G. and R.L.) visually inspected PET images and determined PET-based Braak-like stages.^21^ Visual reads were classified as negative (T-; Braak 0), positive in the medial temporal lobe only (T_+MTL_ or Braak I-II), or positive in the neocortex (T_+NEO_ or Braak ≥ 3 depending on the affected brain regions). For participants without tau-PET data (mostly from Paris and Bordeaux), a CSF p-tau181 concentration ≥60pg/mL defined T+ status.^22^

### Plasma assays

Blood was collected in K_2_-EDTA tubes, independently of fasting status, centrifuged at room temperature at 2500 g for 10 min and plasma aliquots were frozen at −80°C within 2 hours. Sampling occurred within 12 months of the amyloid-PET scan, CSF lumbar puncture, and tau-PET acquisition. After 1 hour of thawing at room temperature, plasma p-Tau217 was measured locally in each center with the Lumipulse^®^ G pTau217 Plasma RUO assay (Fujirebio, Ghent, Belgium) using three independent Lumipulse analyzers (G600II in Brussels and Bordeaux and G1200 in Paris). Plasma NfL was quantified using the Atellica IM® NfL assay in Brussels (Siemens Healthineers, Erlangen, Germany) or using the Lumipulse^®^ G NfL Blood assay in Bordeaux. In Brussels, plasma p-Tau181, Aβ42 and Aβ40 were measured on a Simoa® SR-X (pTau-181 V2.1; Neurology 3-Plex A), and BD-tau on the Simoa® HD-X (BD-Tau Advantage Plus). In Paris, plasma p-tau181, Aβ42, and Aβ40 were measured on Lumipulse G1200 using corresponding RUO assays.

### Statistical analyses

Analyses were conducted using GraphPad Prism 8. Quantitative variables were summarized as mean±SD or median [IQR] and compared using ANOVA or Kruskal–Wallis tests with appropriate post hoc tests. Categorical variables were compared using χ² tests. Correlations between p-tau217 and other biomarkers or imaging measures were assessed using Spearman’s rank coefficient. In the Brussels cohort, optimal p-tau217 cutoffs for Aβ+ vs Aβ- were determined using Youden’s index. Statistical comparison of the areas under the ROC curves was performed using DeLong’s non-parametric test. Dual-threshold strategies targeting 95% and 97.5% Se/Sp were then defined; values between lower and upper thresholds were considered gray zone. These predefined thresholds were applied without modification to the Paris, Bordeaux, and monogenic FTLD clinical validation cohorts to assess robustness and transferability for predicting amyloid status, and clinical-biological AD.

### Standard Protocol Approvals, Registrations, and Participant Consents

Brussels, Belgium (prospective research cohort): The study was reviewed and approved by the Comité d’éthique hospitalo-facultaire (CEHF), the hospital–faculty ethics committee affiliated with the Cliniques universitaires Saint-Luc and the Université catholique de Louvain (UCLouvain), Brussels, Belgium (protocol UCL-2016-121; registered in the EU clinical trials database European Union Drug Regulating Authorities Clinical Trials Database (EudraCT) 2018-003473-94; ethical approval granted). All participants provided written informed consent.

Bordeaux, France (NEODEM/BioNEODEM): The NEODEM and BioNEODEM studies were reviewed and approved by the Comité de Protection des Personnes (CPP; French regional research ethics committee), CPP Île-de-France VI, France (favorable opinion/ethical approval granted). The CPP registered these protocols under dossier references 60-19 and 74-20, respectively, within the French “Jardé law” framework (Law governing recherches impliquant la personne humaine, i.e., research involving the human person; RIPH), as RIPH category 2 and RIPH category 1 studies, hors produits de santé (HPS, i.e., not primarily evaluating a medicinal product or medical device). All participants provided written informed consent.

Paris, France (routine-care cohort; secondary use of care data/surplus samples): Data and surplus plasma/CSF samples were collected strictly as part of routine clinical care at Assistance Publique–Hôpitaux de Paris (AP-HP), Hôpital Pitié-Salpêtrière, Paris, France. Patients were informed of secondary use and could opt out. Secondary use of routinely collected clinical data and surplus samples was conducted under the Commission nationale de l’informatique et des libertés (CNIL; the French data-protection authority) Reference Methodology MR-004 (MR-004), a standard compliance framework for certain health studies/evaluations based on reuse of existing data. Because this component was considered outside the scope of the Jardé law RIPH framework, formal ethics approval by a CPP or an Institutional Review Board (IRB) was not required (waived). AP-HP acted as the data controller.

Paris, France (monogenic frontotemporal lobar degeneration cohort): Participants provided written consent for secondary use of diagnostic samples and associated data. Data processing complied with CNIL MR-004; no additional CPP/IRB ethics approval was required for this secondary use (waived).

### Data availability

Deidentified data are available upon reasonable request to the corresponding author.

## RESULTS

### Participant Characteristics

Participant characteristics are summarized in **Table 1**.

**Table 1:**
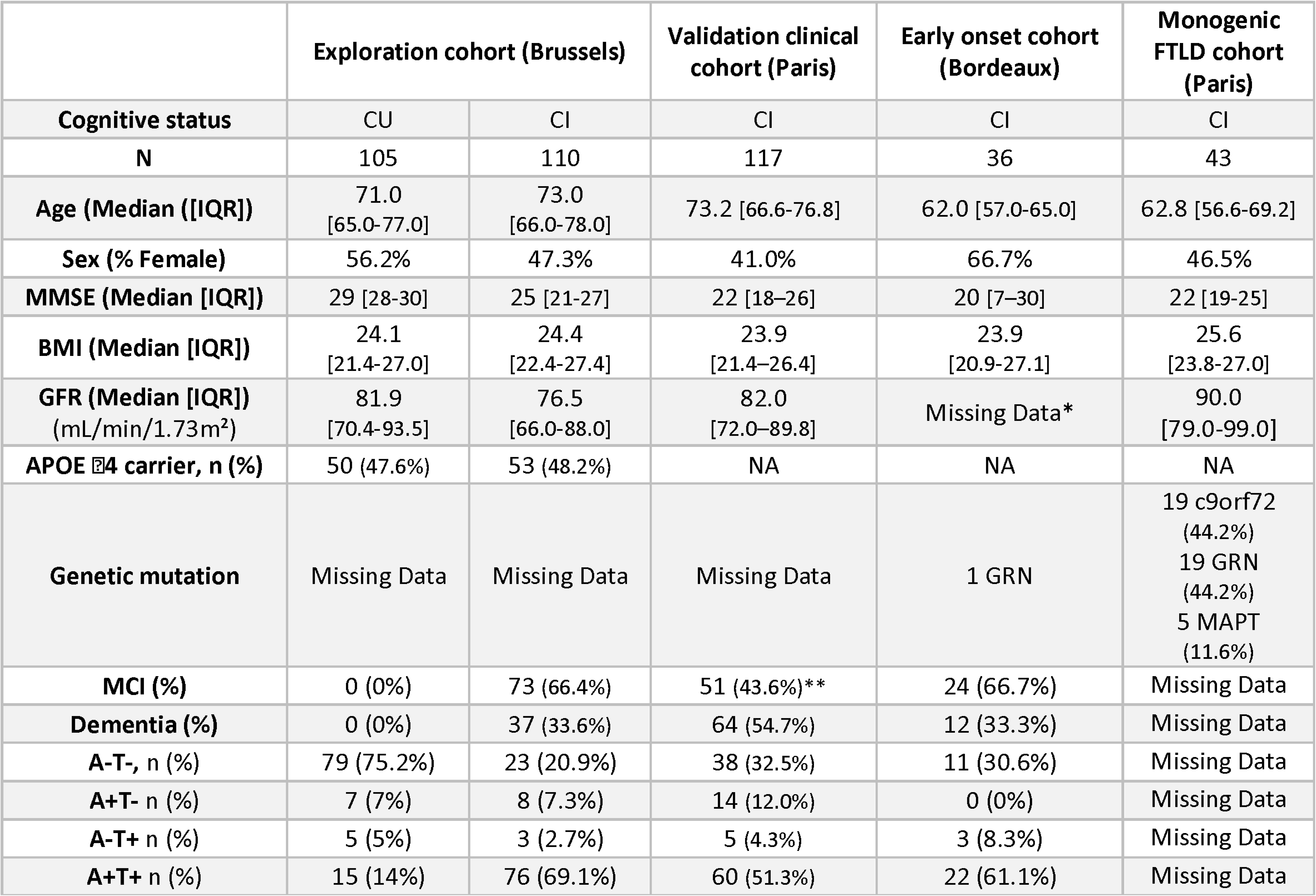
Demographical and clinical characteristics of the study populations. Data are stratified by clinical diagnosis. MMSE = Mini-Mental State Examination; BMI = Body Mass Index; GFR = glomerular filtration rate; IQR = interquartile range; *: GFR was not available in this cohort of young patients, but none of them reported a history of renal impairment. **2 patients of the validation clinical cohort (Paris) were cognitively unimpaired, but attended the memory clinic. NA: Not applicable (APOE l4 testing not routinely analyzed in clinical practice)

The Brussels exploration cohort comprised 215 participants, including 105 cognitively unimpaired (CU; median age 71.0 [IQR 65.0–77.0] years) and 110 cognitively impaired (CI; 73.0 [66.0–78.0] years; 73 MCI and 37 dementia). Women represented 56.2% of CU and 47.3% of CI. CU were enriched for APOE ε4 carriers by design, resulting in comparable ε4-carrier frequencies in CU and CI (∼48%). MMSE was higher in CU than CI (p<0.001), whereas BMI and eGFR were similar between groups (Table 1).

In Paris, 117/131 (89.3%) consecutive memory-clinic patients had paired plasma p-tau217 and CSF biomarkers. Detailed diagnostic distribution is described in **Supplementary Table 2**. Clinical-biological AD was the primary diagnosis in 66/117 (56.4%); other neurodegenerative primary diagnoses in 29/117 (24.8%); and non-neurodegenerative, mixed, or alternative etiologies in 22/117 (18.8%). Comorbid neurodegenerative diagnoses were frequent (24/117, 20.5%). Cognitive status comprised 51/117 (43.6%) MCI, 64/117 (54.7%) dementia, and 2/117 (1.7%) CU (in CU, AD could be considered as a comorbid diagnosis). CSF A/T profiles are provided in Table 1. Amyloid PET (n=3) was used to adjudicate discordant CSF profiles (one positive scan revealing a false-negative A−T− CSF result in typical AD; two negative scans excluding β-amyloidosis). Overall, AD as a primary or comorbid diagnosis was present in 71/117 (60.7%); compared with non-AD, AD patients were slightly older (p=0.03) and had lower MMSE (p<0.001) and BMI (p=0.002), with similar sex distribution and eGFR. Detailed diagnostic distribution in the Paris routine-care cohort are described in Supplementary Table 2.

The Bordeaux early-onset cohort included 36 CI patients (median age 62.0 [57.0–65.0] years), mostly EOAD (n=23), with the remainder including bvFTD (n=3), semantic PPA (n=4), non-fluent PPA (n=1), PSP/CBD (n=2), and uncertain non-AD diagnoses (n=3).

The monogenic FTLD cohort included 43 mutation carriers (C9ORF72 n=19, GRN n=19, MAPT n=5), predominantly presenting with bvFTD (n=22), with fewer PPA variants (n=4) and FTLD-ALS–spectrum presentations (n=1); ALS was documented in three C9ORF72 carriers.

### Defining p-tau217 thresholds in the Brussels exploration cohort

#### BBMs concentrations, clinical status, and brain β-amyloidosis

Results are summarized for all biomarkers in **Table 2** and illustrated for p-tau217 in **Figure 1**.

**Figure 1:**
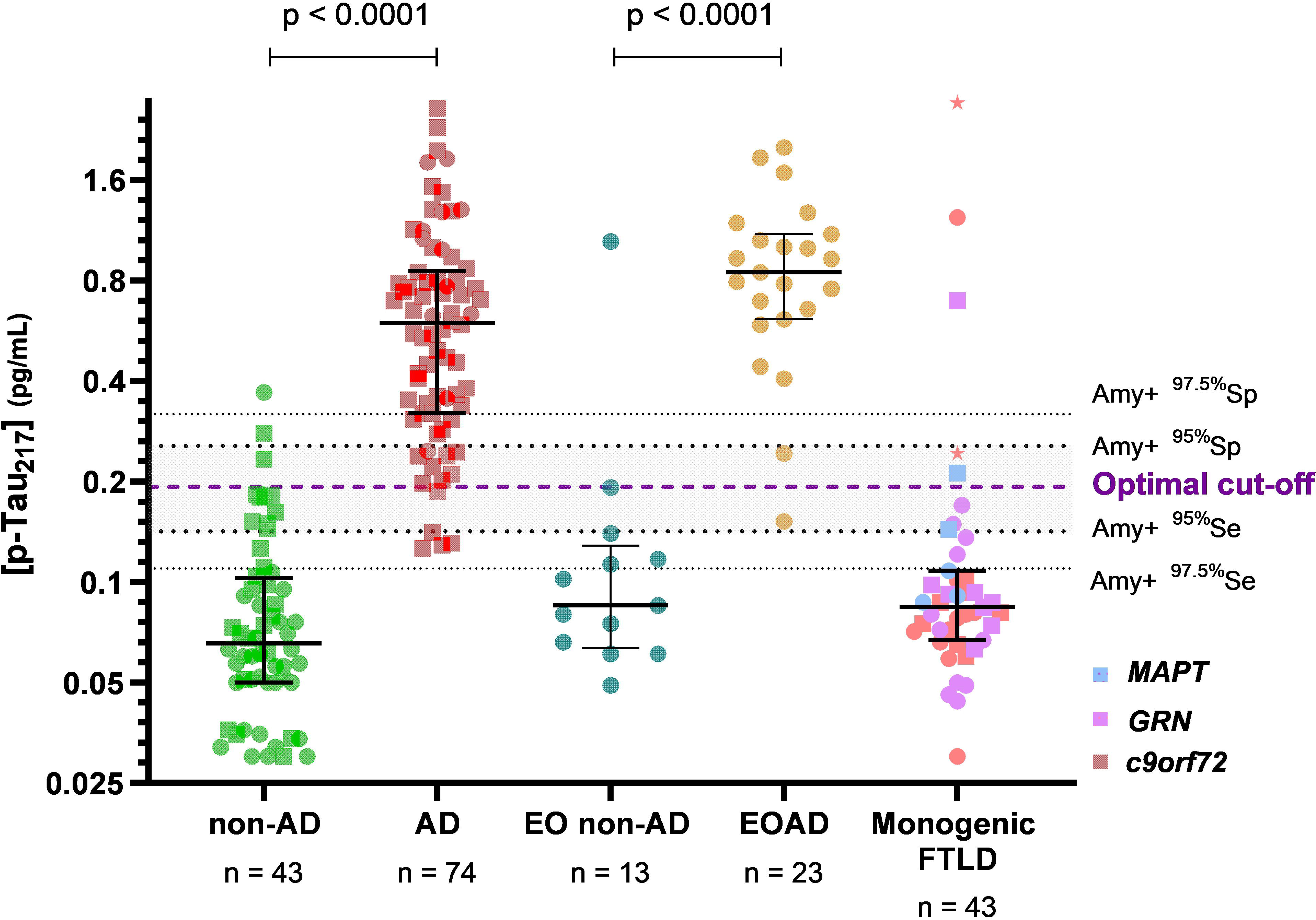
Distribution of plasma p-Tau217 concentrations by amyloid status and Braak stages in the Brussels exploration cohort. Vertical dashed lines indicate thresholds corresponding to 97.5% sensitivity (0.110 pg/mL), 95% sensitivity (0.142 pg/mL) ; optimal cut-off based on the Youden index (0.193 pg/mL ; purple), 95% specificity (0.256 pg/mL) and 97.5% specificity (0.319 pg/mL) for the prediction of Aβ status. The Y-axis is displayed on a log[ scale to improve visualization.

**Table 2:**
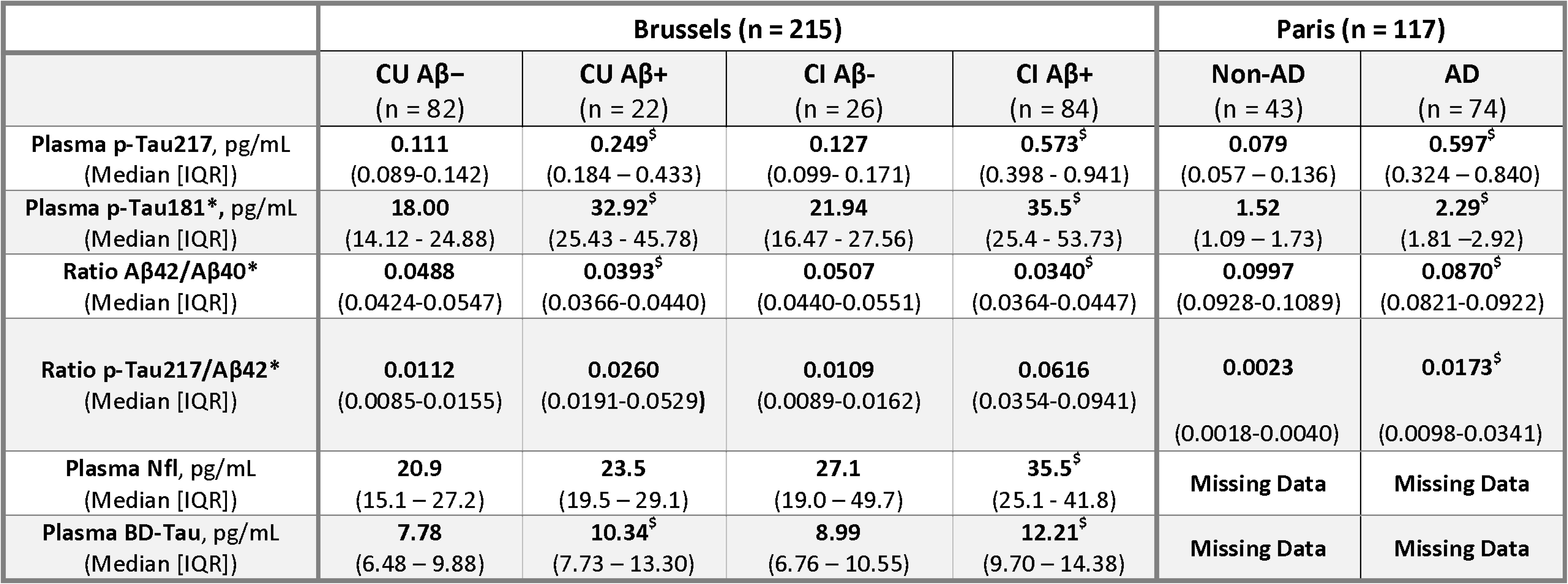
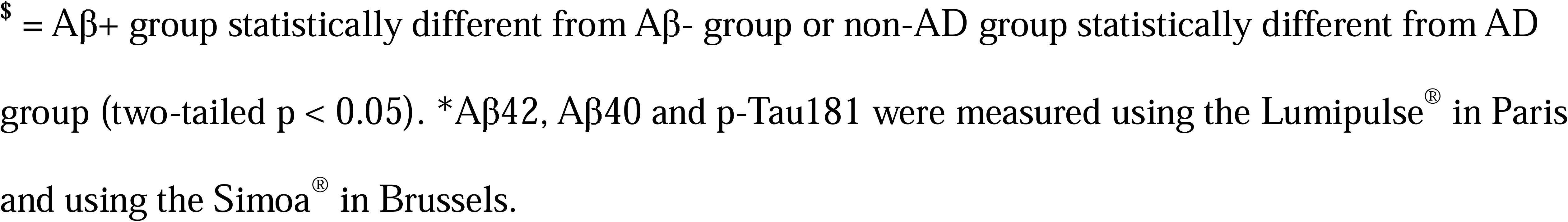
Comparison of plasma biomarker concentrations across clinical and amyloid-defined subgroups. ^$^ =.

|  | Brussels (n = 215) |  |  |  | Paris (n = 117) |  |
| --- | --- | --- | --- | --- | --- | --- |
| | CU Aβ-<br>(n = 82) | CU Aβ+<br>(n = 22) | CI Aβ-<br>(n = 26) | CI Aβ+<br>(n = 84) | Non-AD<br>(n = 43) | AD<br>(n = 74) |
| Plasma p-Tau217, pg/mL<br>(Median [IQR]) | 0.111<br>(0.089-0.142) | 0.249<sup>s</sup><br>(0.184 - 0.433) | 0.127<br>(0.099- 0.171) | 0.573<sup>s</sup><br>(0.398 - 0.941) | 0.079<br>(0.057 - 0.136) | 0.597<sup>s</sup><br>(0.324 - 0.840) |
| Plasma p-Tau181*, pg/mL<br>(Median [IQR]) | 18.00<br>(14.12 - 24.88) | 32.92<sup>s</sup><br>(25.43 - 45.78) | 21.94<br>(16.47 - 27.56) | 35.5<sup>s</sup><br>(25.4 - 53.73) | 1.52<br>(1.09 - 1.73) | 2.29<sup>s</sup><br>(1.81 - 2.92) |
| Ratio Aβ42/Aβ40*<br>(Median [IQR]) | 0.0488<br>(0.0424-0.0547) | 0.0393<sup>s</sup><br>(0.0366-0.0440) | 0.0507<br>(0.0440-0.0551) | 0.0340<sup>s</sup><br>(0.0364-0.0447) | 0.0997<br>(0.0928-0.1089) | 0.0870<sup>s</sup><br>(0.0821-0.0922) |
| Ratio p-Tau217/Aβ42*<br>(Median [IQR]) | 0.0112<br>(0.0085-0.0155) | 0.0260<br>(0.0191-0.0529) | 0.0109<br>(0.0089-0.0162) | 0.0616<br>(0.0354-0.0941) | 0.0023<br>(0.0018-0.0040) | 0.0173<sup>s</sup><br>(0.0098-0.0341) |
| Plasma NfL, pg/mL<br>(Median [IQR]) | 20.9<br>(15.1 - 27.2) | 23.5<br>(19.5 - 29.1) | 27.1<br>(19.0 - 49.7) | 35.5<sup>s</sup><br>(25.1 - 41.8) | Missing Data | Missing Data |
| Plasma BD-Tau, pg/mL<br>(Median [IQR]) | 7.78<br>(6.48 - 9.88) | 10.34<sup>s</sup><br>(7.73 - 13.30) | 8.99<br>(6.76 - 10.55) | 12.21<sup>s</sup><br>(9.70 - 14.38) | Missing Data | Missing Data |

\$ = Aβ + group statistically different from Aβ - group or non-AD group statistically different from AD group (two-tailed p < 0.05). \*Aβ42, Aβ40 and p-Tau181 were measured using the Lumipulse® in Paris and using the Simoa® in Brussels.

Amyloid status was determined using PET in 124 participants (95 CU, 22 MCI, 7 DEM) and by CSF Aβ42 in 91 (10 CU, 51 MCI, 30 DEM). Overall, 109/215 (51%) were Aβ+: 21% of CU, 66% of MCI, and 97% of DEM.

Across cognitive strata, Aβ+ individuals had markedly higher p-tau217 than Aβ- individuals (0.503 [0.297-0.858] vs 0.112pg/mL [0.091-0.149], p<0.0001). In detail, median p-tau217 was 0.249pg/mL [0.176–0.452] in Aβ+ CU and 0.111pg/mL [0.088–0.141] in Aβ- CU; while median p-tau217 concentration reached 0.573pg/mL [0.398–0.941] in Aβ+ CI versus 0.127pg/mL [0.100–0.168] in Aβ- CI (all p<0.0001). Plasma p-tau217 values did not significantly differ between CU Aβ- and CI Aβ-. In the entire cohort, p-tau217 discriminated Aβ+ and Aβ- individuals with an AUC of 0.96 (95% CI 0.93–0.98). The Youden cutoff at 0.193pg/mL yielded 91.5% sensitivity and 91.6% specificity. Dual 95% Se/Sp thresholds (0.142/0.256 pg/mL) defined a gray zone of 19.7%; Dual 97.5% Se/Sp thresholds (0.110/0.319pg/mL) expanded it to 39.2% (**Table 3**). Restricting the data to A+T+ versus A−T− individuals yielded an AUC of 0.98 with the same optimal cutoff (0.193 pg/mL).

**Table 3:** Diagnostic performance of BBMs for predicting amyloid pathology in the Brussels exploration cohort. Additional thresholds corresponding to 95% and 97.5% sensitivity (Se) and specificity (Sp) are also shown. The percentage of individuals falling into the “gray zone” between the 95% Se and 95% Sp thresholds is indicated (^1^), as well as between the 97.5% Se and 97.5% Sp thresholds (^2^). NEG = proportion of subjects between 95%Se cutoff and the Youden cutoff (^1^) or between the 97.5%Se cutoff and the Youden cutoff (^2^). POS = proportion of subjects between Youden cutoff and 95%Sp cutoff (^1^) or between the Youden cutoff and 97.5%Sp cutoff (^2^).

| Biomarker | Youden Cutoff | Sensitivity (CI<sub>95</sub>) | Specificity (CI<sub>95</sub>) | 95% Se cutoff | 95% Sp cutoff | 95% gray zone (NEG & POS)<sup>1</sup> | 97.5% Se cutoff | 97.5% Sp cutoff | 97.5% gray zone (NEG & POS)<sup>1</sup> | AUC ROC |
| --- | --- | --- | --- | --- | --- | --- | --- | --- | --- | --- |
| p-Tau217 (pg/mL) | 0.193 | 91.5%<br>(IC95: 84.7-95.5%) | 91.6%<br>(IC95: 84.8-95.5%) | 0.142 | 0.256 | 19.7%<br>(11.7 & 8.0%) | 0.110 | 0.319 | 37.2%<br>(25.6 & 11.6%) | 0.959 |
| AB42/AB40 | 0.0465 | 85.1%<br>(IC95: 76.6-90.9%) | 61.9%<br>(IC95: 51.9-70.9%) | 0.0523 | 0.0349 | 68.4%<br>(18.1 & 50.3%) | 0.0708 | 0.0267 | 93.8%<br>(58.0 & 35.8%) | 0.787 |
| p-Tau181 (pg/mL) | 24.96 | 76.7%<br>(IC95: 67.0-84.2%) | 75.8%<br>(IC95: 66.3-83.3%) | 16.02 | 45.15 | 60.0%<br>(29.7 & 30.3%) | 12.00 | 53.44 | 80.2%<br>(43.9 & 45.3%) | 0.812 |
| p-Tau217/AB42 | 0.0184 | 88.3%<br>(IC95: 80.3-93.3%) | 88.7%<br>(IC95: 80.8-93.6%) | 0.0104 | 0.0258 | 35.1%<br>(26.2 & 8.9%) | 0.0095 | 0.0263 | 40.4%<br>(31.1 & 9.3%) | 0.941 |
| p-Tau217/AB42/AB40 | 3.900 | 90.4%<br>(IC95: 82.8-94.9%) | 86.6%<br>(IC95: 78.4-92.0%) | 2.579 | 6.155 | 25.7%<br>(14.1 & 11.5%) | 2.014 | 6.750 | 40.4%<br>(27.4 & 13.0%) | 0.941 |
| NfL (pg/mL) | 26.6 | 65.1%<br>(IC95: 55.6-73.5%) | 69.2%<br>(IC95: 59.9-77.1%) | 15.7 | 93.8 | 82.2%<br>(37.6 & 44.6%) | 13.6 | 104.0 | 90.7%<br>(44.2 & 46.5%) | 0.694 |
| BD-Tau (pg/mL) | 9.99 | 68.4%<br>(IC95: 58.5-76.7%) | 76.4%<br>(IC95: 66.8-84.4%) | 6.44 | 15.46 | 72.4%<br>(39.8 & 32.6%) | 5.38 | 16.78 | 82.0%<br>(47.0 & 35.0%) | 0.774 |

P-tau217 significantly outperformed Aβ42/Aβ40, p-tau181, BD-tau, NfL, and all composite ratios for predicting Aβ status (DeLong p<0.0001). The p-tau217/Aβ42 and p-tau217/Aβ42/Aβ40 ratios (AUCs = 0.94) did not improve accuracy or reduce gray zones. In participants with amyloid-PET, p-tau217 correlated with Centiloid values (Spearman ρ=0.63), more strongly than with BD-tau (ρ=0.42), p-tau181 (ρ=0.40), or Aβ42/Aβ40 (ρ=−0.29, p<0.001); however, NfL showed no significant correlation with Centiloid values. P-tau217 correlated inversely with eGFR in the overall sample (ρ=−0.26, p<0.0001) and in Aβ-(ρ=−0.25, p=0.011), but did not reach statistical significance in Aβ+ (ρ=−0.16, p=0.10). No association with BMI was observed, either in the overall cohort or within the Aβ− and Aβ+ subgroups.

#### BBMs and tauopathy

Among 185 participants with tau-PET, T+ individuals (T_+MTL_ or T_+NEO_) had 4.34-fold higher median p-tau217 than T- individuals (0.503 [0.290-0.851] vs 0.116pg/mL [0.090-0.167], p<0.0001). P-tau217 discriminated T+ from T- with an AUC of 0.93 (95% CI 0.90–0.97); the amyloid-anchored p-tau217 0.193pg/mL cutoff yielded 89.0% sensitivity and 86.3% specificity, indicating that some A+T- individuals were above that threshold. P-tau217 correlated strongly with temporal meta-ROI tau-PET SUVR (ρ=0.76, p<0.0001), while correlations were weaker for Aβ42/Aβ40, NfL, and BD-tau. Median p-tau217 rose with PET-based Braak stages (**Figure 2)**: 0.114 [0.088–0.167] (stage 0), 0.167 [0.108–0.222] (I–II), 0.435 [0.218–0.550] (III–IV), 0.649 [0.450–1.006] pg/mL (V–VI).

**Figure 2.**
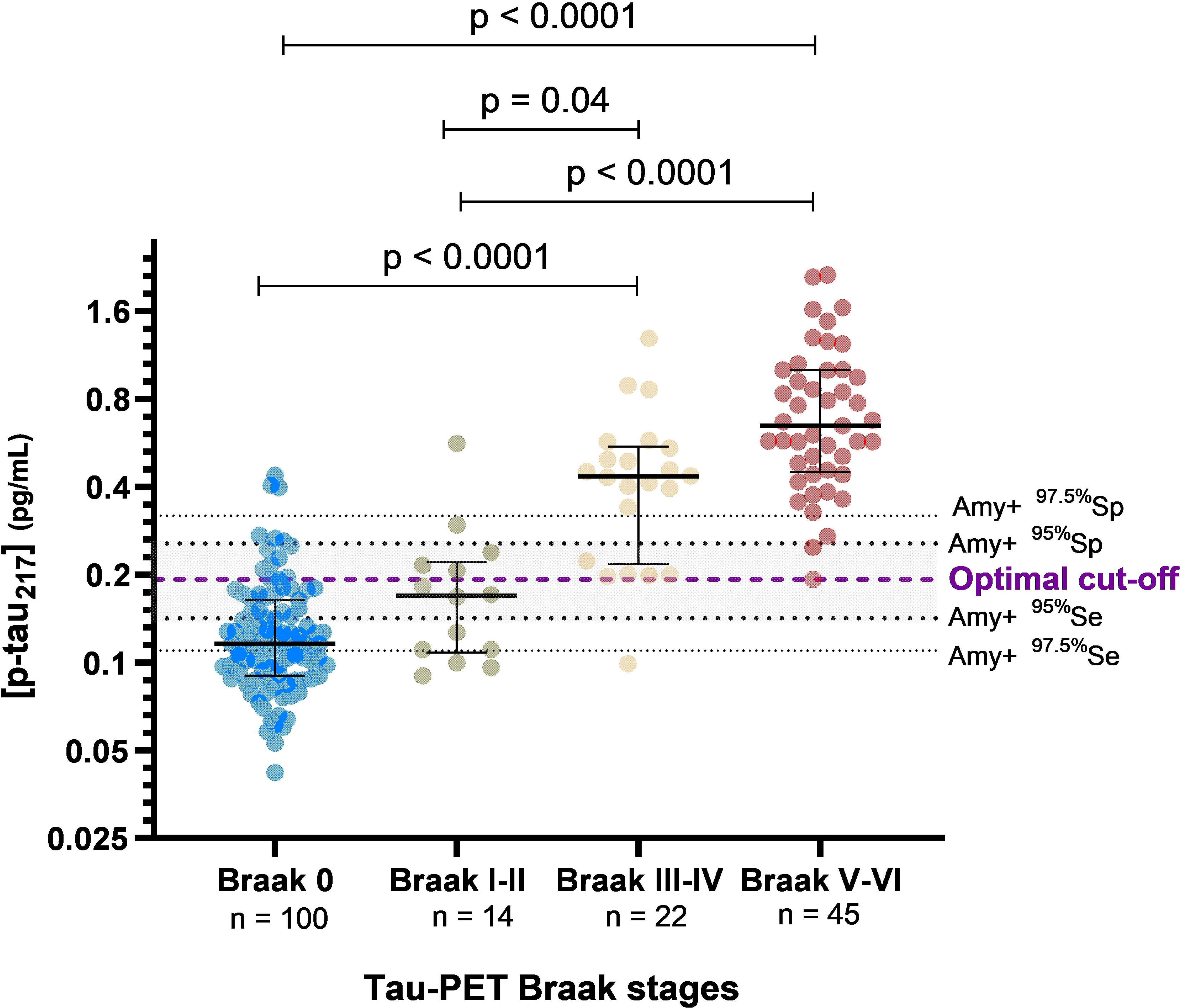
: Plasma p-Tau217 concentrations across Tau-PET Braak stages. Bars represent median concentrations ± IQR. Tau-PET positivity in the MTL and neocortex and in each Braak region was determined using visual assessment of two trained nuclear physicians. The Y-axis is displayed on a logl scale to improve visualization

#### Diagnostic performances of p-Tau217 thresholds for predicting Tau-PET status

Using amyloid-anchored thresholds, 0.193pg/mL detected T+_MTL_ with 89.0% sensitivity and 86.3% specificity and T_+NEO_ with 98.5% sensitivity and 81.8% specificity, indicating that concentrations below 0.193 pg/mL reliably exclude the presence of neocortical tauopathy. Using 95% Se/Sp dual thresholds, sensitivity/specificity for T+_MTL_ were 92.6%/92.7% and for T_+NEO_ 98.5%/90.9% (21.0% gray zone). Using 97.5% dual thresholds, sensitivity/specificity for T_+MTL_ were 95.1%/95.8% and for T_+NEO_ 98.5%/94.5% (39.2% gray zone) (**Table 4**).

**Table 4:**
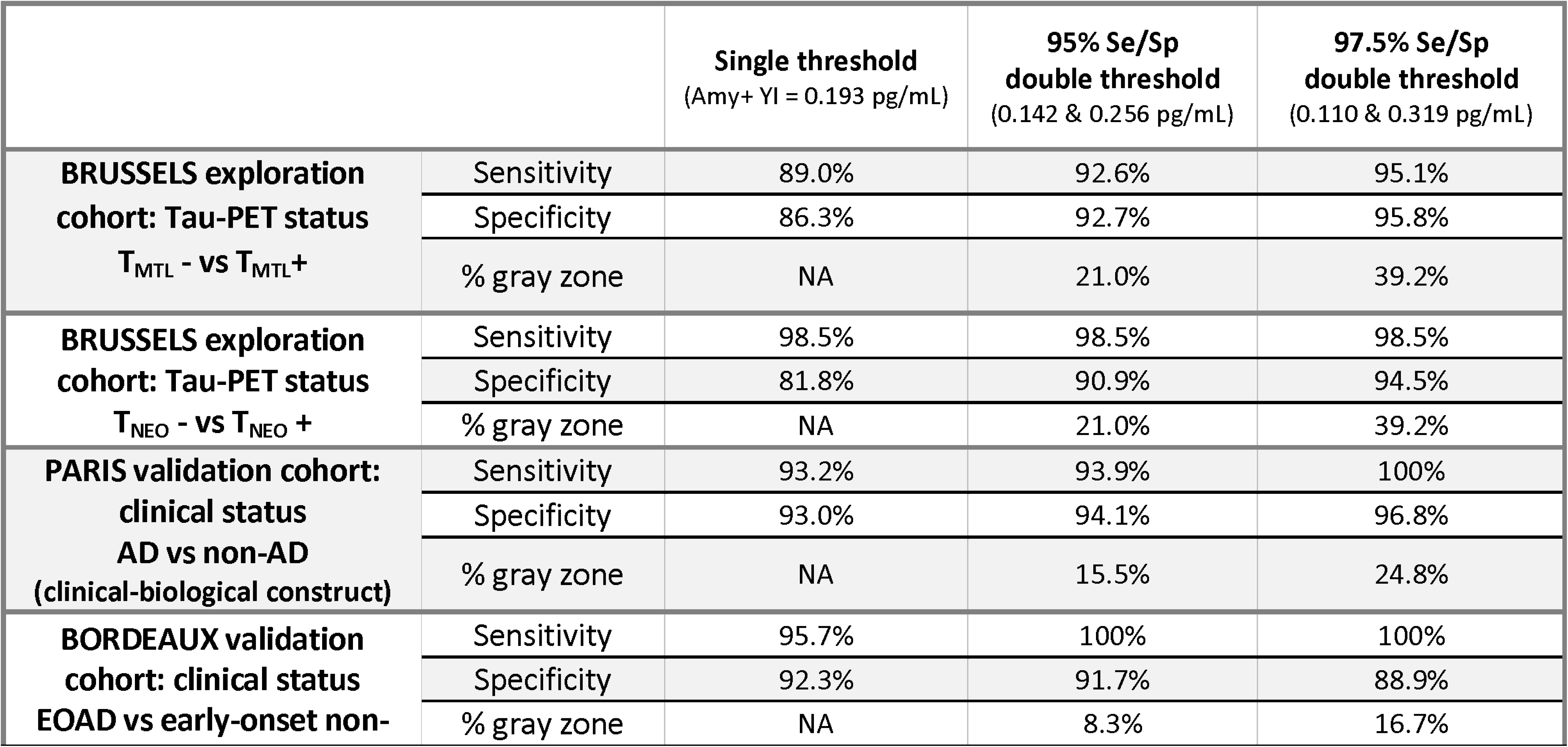
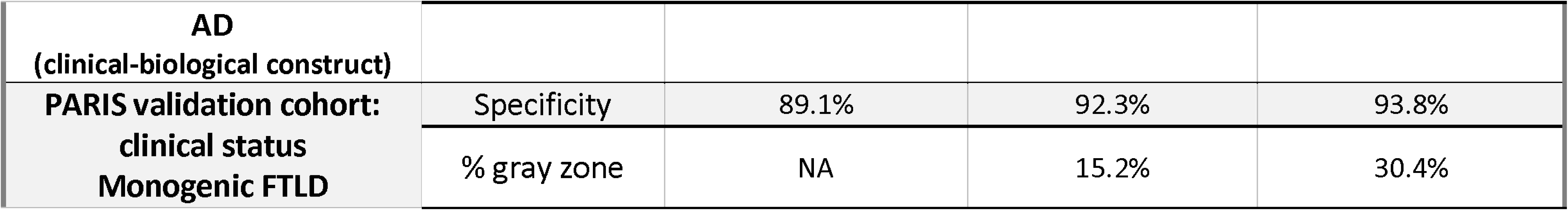
Diagnostic performance of predefined plasma p-Tau217 thresholds for predicting amyloid and tau pathology across independent cohorts. Diagnostic performance for identification of tau-PET positivity (line 1 and 2). Performances are shown separately for two tau-PET classification systems: T_MTL+_ (medial temporal tau deposition) and T_NEO+_ (neocortical tau deposition). Line 3 and 4 : Diagnostic performance for identification of AD and EOAD in the Paris and Bordeaux cohorts, respectively.

### External validation of predefined p-tau217 thresholds

External validation is provided in **Table 4** and illustrated in **Figure 3**.

**Figure 3:**
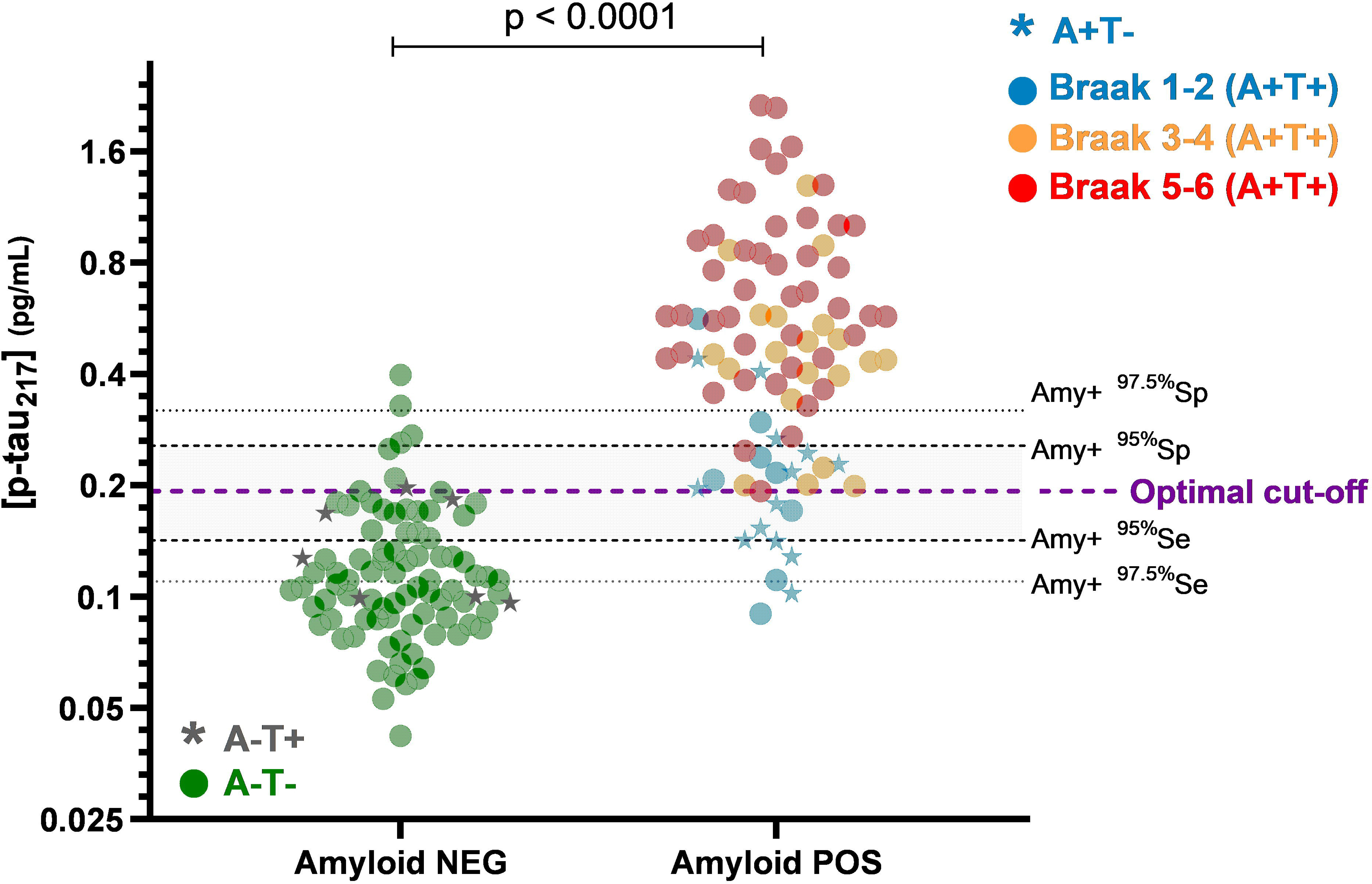
p-Tau217 concentrations measured in the external validation cohorts. p-Tau217 values are issued from the Paris cohort (non-AD and AD), the Bordeaux’s cohort (Early-onset AD and non-AD) as well as in the Paris Monogenic FTLD cohort. Square: patients > 65 years; circle: patients ≤ 65 years; star: patients with a defined clinical and electrophysiological amyotrophic lateral sclerosis (ALS). The Y-axis is displayed on a log[ scale to improve visualization.

#### Paris memory clinic cohort

Plasma p-tau217 was substantially higher in AD than non-AD patients (0.597 [0.324–0.840] vs 0.079 [0.057–0.136] pg/mL). The local Youden cutoff (0.185 pg/mL) yielded 94.6% sensitivity and 93.0% specificity (AUC 0.975, **Supplementary Table 1** for detailed Paris data). The Brussels amyloid-anchored 0.193 pg/mL cutoff achieved 93.2% sensitivity, 93.0% specificity, PPV 95.7%, NPV 88.5%.

Using the 95% Se/Sp dual thresholds defined in Brussels (0.142/0.256pg/mL), we obtained 93.9% sensitivity, 94.1% specificity, PPV 96.5%, NPV 90.0%, and a 15.4% gray zone. With 97.5% dual thresholds (0.110/0.319 pg/mL), sensitivity reached 100%, specificity 96.8%, PPV 98.3%, NPV 100%, with a 24.8% gray zone (58 positive, 30 negative, 29 gray). Gray-zone resolution by CSF as second-line biomarker testing was high. With 97.5% dual thresholds, 22/28 (78.6%) gray-zone patients had unambiguous A+T+ or A−T− CSF profiles; with 95% thresholds, 13/18 (76.5%) gray-zone patients had A+T+ or A−T− (**Supplementary Tables 3and 4**).

Plasma p-tau217 did not significantly correlate with eGFR in the full cohort or AD subgroup; an inverse trend in non-AD did not reach significance (ρ=−0.28, p=0.08). A modest inverse correlation with BMI in the full cohort disappeared after stratification by AD status (whole cohort: ρ=−0.35, p<0.001; AD: ρ=−0.17, p = 0.17); non-AD: ρ=−0.20, p=0.22). No other plasma biomarker or ratio outperformed p-tau217 or reduced gray-zone proportions (**Supplementary Table 1**).

Three non-AD participants had plasma p-tau217 >0.193 pg/mL (7.0%). These included an individual aged 80–84 years with idiopathic normal pressure hydrocephalus and normal CSF AD biomarkers; an individual aged 70–74 years with subjective cognitive complaints/anxiety and normal CSF AD biomarkers; and an individual aged 55–59 years with bvFTD in whom an FTLD–ALS spectrum process was clinically suspected despite non-confirmatory electrophysiology. To protect confidentiality, additional individual-level details are not reported. Among AD patients (n=74), only 5 (6.8%) had plasma p-tau217 in the low 97.5% Se/Sp gray zone (0.110–0.193 pg/mL). All had mild disease (MCI; MMSE 26-29) with CSF profiles fully consistent with AD. Their clinical phenotypes were amnestic in 4/5 (with secondary dysexecutive in 2/5 and language impairment in 1/5) and language-led in 1/5. One-year follow-up in 2025 confirmed progression compatible with clinical-biological AD. FDG-PET was available for 4/5, showing either normal neocortical metabolism (2/4), isolated bilateral hippocampal hypometabolism without posterior involvement (1/4), or a more AD-like pattern with left parietal hypometabolism and bilateral prefrontal involvement, while posterior cingulate/precuneus remained preserved (1/4).

#### Bordeaux early-onset cohort

In the early-onset dementia cohort, the 0.193pg/mL cutoff discriminated EOAD from non-AD with 95.7% sensitivity and 92.3% specificity (AUC 0.94). The 95% Se/Sp dual thresholds achieved 100% sensitivity, 91.7% specificity, with an 8.3% gray zone; 97.5% thresholds provided 100% sensitivity, 88.9% specificity, with a 16.7% gray zone (**Table 4**).

A patient with a clinical presentation of bvFTD and brain imaging (MRI and FDG-PET) highly suggestive of this diagnosis had a very high p-tau217 value (1.049pg/mL), despite an A-/T- profile in the CSF that was well above the established thresholds. Their blood NfL level was 178.6 pg/mL, which is a level compatible with ALS. However, one year after sampling, there were no clinical or electromyographic findings suggestive of lower motor neuron involvement. The patient had no family history of FTD or ALS and chose not to undergo *C9ORF72* testing.

#### Monogenic FTLD cohort

In the monogenic FTLD cohort (all considered non-AD), the 0.193 pg/mL cutoff yielded 89.1% specificity (5/43 >0.193 pg/mL, **Table 4**). Applying the 95% Se/Sp dual thresholds increased specificity to 92.3% (15.2% gray zone), and the 97.5% thresholds to 93.8% (30.4% gray zone). Amongst the five patients with p-tau217 > 0.193 pg/mL, three carried *C9ORF72* expansion: two had EMG-confirmed ALS with combined cognitive-behavioral and lower motor neuron involvement, and one had bvFTD with upper motor neuron signs; clinical follow-up information was limited. The remaining two were older mutation carriers (GRN, age 75–79 years; MAPT, age 65–69 years) with elevated p-tau217 and limited AD biomarker work-up, in whom AD co-pathology could not be reliably excluded.

## DISCUSSION

This multicenter study demonstrates that plasma p-tau217 measured on the Lumipulse platform provides robust, transferable diagnostic thresholds for identifying Alzheimer’s pathology across heterogeneous populations and laboratory environments. Thresholds defined in a single research cohort generalized with high accuracy to routine memory clinic, early-onset neurodegenerative dementia, and monogenic FTLD cohorts, supporting harmonized use in clinical practice. Moreover, we emphasize the usefulness of CSF as a secondary biomarker investigation for individuals in the gray zone after plasma testing, and the appropriateness of plasma p-tau217 as a standalone biomarker when values exceed a high threshold (e.g., 0.319pg/mL for Sp=97.5%), including for initiating amyloid-lowering therapies.^27^

Plasma p-tau217 consistently outperformed other BBMs, including Aβ42/Aβ40, p-tau181, NfL, BD-tau, and composite ratios, in predicting Aβ status and distinguishing clinical-biological AD from non-AD, aligning with previous reports.^7,28,29^ Unlike Lehmann and colleagues., the p-tau217/Aβ42 ratio did not improve diagnostic accuracy or reduce the gray zone in the exploration (Brussels) or validation (Paris) cohorts, despite using distinct assays for Aβ42 measurement.^10^ This may be due to Lehmann et al. using 90% Se/Sp dual-cutoff strategies in research cohorts, which may have corrected variability and compressed the gray zone, unlike our stricter 95 or 97.5% Se/Sp thresholds and the low prevalence of participants with eGFR < 45 mL/min in the present study, resulting in a narrower gray zone and limited benefit from the ratio. However, p-tau217 and Aβ42 exhibit markedly different pre-analytical stability profiles.^30,31^ Therefore, ratio-based measures such as p-tau217/Aβ42 may be disproportionately affected by pre-analytical handling conditions, potentially leading to artifactual ratio alterations when samples are not promptly frozen; this effect may be particularly pronounced under real-world routine conditions, where pre-analytical constraints are inherently more variable.^30^ These results support p-tau217 as a stand-alone primary BBM on Lumipulse in similar clinical settings, while acknowledging that ratio approaches may remain useful in cohorts with higher proportion of comorbidities, like chronic kidney disease.

Established cutoffs reliably detected amyloid and clinical-biological AD across diverse cohorts and settings, supporting their transferability. Consistent performance across independent analyzers, clinical phenotypes (including late-onset and early onset symptomatic AD, and non-AD neurodegenerative disorders), and both research and routine care attests to the robustness of the Lumipulse p-tau217 assay. Importantly, our 95% Se/Sp thresholds (0.142–0.256 pg/mL) align with recent estimates of the Alzheimer’s Disease Neuroimaging Initiative (ADNI) (0.164-0.292 pg/mL), which were derived using a modelling approach based on published data ^32^. Diagnostic accuracy was over 90% across four settings, with the lowest in the monogenic FTLD cohort, where the proportion of lower motor neuron involvement was also higher. Applying the research thresholds (95% and 97.5%) yielded more than 93% accuracy in clinics. Sensitivity reached 100% in Paris and Bordeaux, and gray zones remained below 40%. In a routine clinical care setting, the gray zone was typically around 25%, and under 16% with the 95% thresholds. These gray zones balance clinical practicality and robust diagnosis performance. This also conforms to existing literature, where 95% dual thresholds generally result in gray zones of approximately 12–30%, whereas 97.5% dual threshold approaches tend to expand gray zones to about 40–60% or more.^6,33–35^

In the Brussels research setting, p-tau217 alone showed less sensitivity to early tau deposition (T_MTL+_) detected by tau-PET, indicating a diagnostic gap for initial tau pathology detection. These results are in line with previous published data.^36–38^ Alternatively, early tau deposition—when measured with a second-generation tau-PET tracer—may lack sensitivity for intermediate Alzheimer’s pathology levels that could explain prodromal clinical-biological AD, especially given the high proportion of Tau PET Braak stages I-II in the plasma pTau217 gray zone (**Figure 1**). In such cases, using pTau217 as an earlier marker, i.e., anchored at amyloid PET positivity, might outperform Tau PET to highlight prodromal AD.

The cases of p-tau217 elevations in non-AD patients further refine test interpretation. Across the Paris, Bordeaux, and monogenic FTLD cohorts (n=196), virtually all non-AD individuals with p-tau217 above the amyloid-positivity threshold could be attributed either to FTLD–ALS-spectrum disease with lower motor neuron involvement or to plausible comorbid or clinically-silent Alzheimer’s pathology, consistent with recent evidence of p-tau217 alterations in ALS.^39^ Only three cases (1.5%) were left unexplained, as these samples could not be retested. Thus, true unexplained false positivity appears rare; markedly elevated p-tau217 in non-AD contexts should prompt systematic evaluation for kidney dysfunction, motor neuron disease, and AD co-pathology.^39,40^

Our results inform threshold selection for different clinical scenarios. In routine memory clinics, a single cutoff of 0.193 pg/mL may be more practical, balancing sensitivity and specificity, supporting decision-making, and accounting for real-world constraints such as limited access to confirmatory imaging or constraints related to lumbar puncture eligibility. In this setting, the NPV was 88.9%, and the PPV was 95.8% from the Paris clinical routine cohort. This indicates that in routine care, about 1 in 10 patients could be wrongly excluded for AD after a negative plasma pTau217 test, and 1 in 25 might be improperly diagnosed with AD after a positive result. In the Brussels exploration cohort, the 0.193 pg/mL threshold also provided the best trade-off between sensitivity and specificity for highlighting brain β-amyloidosis or T_MTL+_ and excluding neocortical tau (T_NEO+_). Therefore, patients misclassified as AD-negative using this cutoff most likely do not have neocortical tauopathy, as shown by tau-PET imaging. This may still indicate intermediate levels of Alzheimer’s pathology in a patient with prodromal AD, but is more likely to reflect comorbid Alzheimer’s pathology in patients with more severe impairment. It may offer a pragmatic compromise, accepting that a small minority—often with mild or comorbid pathology—may be misclassified.

On the other hand, our results provide empirical support for a pragmatic two-step diagnostic algorithm, a strategy increasingly relevant with the introduction of amyloid-lowering therapies. Plasma p-tau217 is used as the first-line test, and CSF (or PET where appropriate) is reserved for individuals in the plasma-defined gray zone or with discordant clinical-biological profiles. In the memory clinic cohort from Paris, dual 95% and 97.5% thresholds constrained gray zones to ∼16% and ∼25%, respectively, and, within these subsets, CSF A/T status was unambiguous (A+T+ or A−T−) in about three-quarters of patients, confirming that targeted CSF testing efficiently resolves most indeterminate plasma results. Only a minority with discordant A+T− or A−T+ required additional integrative assessment. These findings operationalize conceptual proposals based on p-tau217 dual thresholds and show that a limited second-line CSF strategy can maintain high diagnostic certainty while minimizing invasive procedures compared with current plasma-free AD biomarker investigations.^6,33–35^ Furthermore, the 97.5% Se/Sp dual threshold yields 100% NPV and 98.3% PPV in Paris’ validation cohort and could serve as a rule-out criterion, as p-tau217 below this level suggests minimal amyloid or tau pathology, mostly precluding the need for confirmatory CSF or PET tests. It may also guide therapy decisions, especially for anti-amyloid treatments, with high PPV supporting treatment initiation without further tests after excluding lower motor neuron involvement or renal impairment.^27^ The gray zone of about 25% means it still reduces the need for biomarker tests compared to current plasma-free approaches. A 95% Se/Sp scheme offers an intermediate balance with smaller gray zones and excellent accuracy, still in line with the recommendations from the global CEO Initiative on AD.^41^

Limitations include recruitment from specialized centers and a majority of symptomatic individuals, limiting extrapolation to preclinical or population screening, where lower pretest probabilities reduce PPV.^42^ In line with IWG recommendations and recent experimental data, p-tau217 should be interpreted within a clinical-biological framework, not as a stand-alone screening test in CU individuals^43,44^. Individuals with severe renal dysfunction were underrepresented; dedicated studies are needed in this group. Finally, while our multi-cohort, multi-analyzer design strengthens generalizability, further validation across additional laboratories, ancestries, and assay lots within formal standardization efforts remains necessary to define universal decision limits.

Altogether, our multicenter data support the implementation of plasma p-tau217, measured on the Lumipulse platform, as a robust first-line biomarker for identifying AD pathology across heterogeneous clinical and laboratory settings. A single threshold of 0.193 pg/mL is proposed for diagnostic purposes without treatment implications, acknowledging that a small minority (5 to 10%) —often with mild or comorbid pathology— may be misclassified. In the context of upcoming anti-amyloid therapies or in patients with discordant clinical-biological profiles, a pragmatic two-step strategy, reserving CSF or PET biomarkers for patients in predefined gray zones, can ensure >95% diagnostic certainty while limiting invasive or costly procedures.

## FUNDING AND DISCLOSURES/CONFLICT OF INTEREST

This study was **not** funded by Fujirebio.

The Brussels Tau-PET study (UCL-2016-121) has been funded thanks to the StopAlzheimer Foundation (PI: Hanseeuw, 2022-2027) as well as a WelBio Starting Grant from the WEL Research Institute (PI: Hanseeuw, 2022-2026).

The BioNEODEM biobank, an ancillary study of the NEODEM cohort (early-onset dementias in Bordeaux), was funded by the French Rotary Club; the measurement of pTau-217 was funded by Lions Alzheimer.

J.L.B received speaker fees from Siemens Healthineers, The Binding Site Company and Fujirebio, outside the submitted work.

Independent of this work, NV received research support from Fondation Bettencourt-Schueller, Fondation Servier, Union Nationale pour les Intérêts de la Médecine (UNIM), Fondation Claude Pompidou, Fondation Alzheimer, Banque Publique d’Investissement, Lion’s Club Alzheimer and Fondation pour la Recherche sur l’Alzheimer; travel grant from the

Movement Disorders Society, Merz-Pharma, UCB Pharma, and GE Healthcare SAS; is an unpaid local principal investigator or sub-investigator in NCT05531526 (AR1001, AriBio), NCT06079190 (AL101, GSK), NCT04241068 and NCT05310071 (aducanumab, Biogen), NCT05399888 (BIIB080, Biogen), NCT03352557 (gosuranemab, Biogen), NCT04592341 (gantenerumab, Roche), NCT03887455 (lecanemab, Eisai), NCT03828747 and NCT03289143 (semorinemab, Roche), NCT04619420 (JNJ-63733657, Janssen – Johnson & Johnson), NCT06544616 (JNJ-64042056, Janssen – Johnson & Johnson), NCT04374136 (AL001, Alector), NCT04592874 (AL002, Alector), NCT04867616 (bepranemab, UCB Pharma), NCT04777396 and NCT04777409 (semaglutide, Novo Nordisk), NCT05469360 (NIO752, Novartis), NCT06647498 (remternetug, Washington University School of Medicine); is the unpaid French national coordinator in NCT05564169 (VHB937, Novartis); has given unpaid lectures in symposia organized by Eisai and the Servier Foundation; has been an unpaid expert for Janssen – Johnson & Johnson, Eli-Lilly, Novartis.

During the past three years, V.P. was a local unpaid investigator or sub-investigator for clinical trials granted by Novo Nordisk, Janssen, Alector, BMS and Roche.

During the past three years, SB was investigator without personal fees in Alzheimer Disease therapeutical trials from Biogen, Roche, Eisai, Eli Lilly, Janssen, Johnson & Johnson, Alector, UCB, NovoNordisk, Novartis, GlaxoSmithKline, and expert without personal fees for GE-Healthcare and Eli-Lilly, all outside the submitted work.

B.J.H. has been a consultant for Biogen, Eisai and Lilly, has received fees as a speaker from Roche over the past five years.

## ACKNOWLEDGMENTS

The authors thank the studies and participants who provided biological samples, the laboratory staff as well as the nurses who recruited and sampled the patients.

## AUTHOR CONTRIBUTIONS

Mr. Bayart: drafting and revising the manuscript for content, study concept and design, analysis and interpretation of data, laboratory analysis, statistical analysis, study supervision, and coordination. Dr. Villain: drafting and revising the manuscript for content, study concept and design, interpretation of data, acquisition of clinical data, statistical analysis, study supervision and coordination. Dr. Planche: drafting and revising the manuscript for content, interpretation of data, acquisition of clinical data. Dr. Boyer: revising the manuscript for content, analysis and interpretation of data. Dr. Colmant: revising the manuscript for content, analysis and interpretation of data. Dr. Le Ber: revising the manuscript for content, interpretation of data, acquisition of clinical data. Dr. Clot: revising the manuscript for content, interpretation of data, acquisition of clinical data. Dr. Bombois: revising the manuscript for content, interpretation of data, acquisition of clinical data. Dr. El-Mazria: revising the manuscript for content, interpretation of data, acquisition of clinical data. Ms. Dingeo: revising the manuscript for content, sample processing, laboratory analysis and acquisition of laboratory data. Ms. Bahroun: revising the manuscript for content, sample processing and acquisition of laboratory data. Dr. Nabeebaccus: revising the manuscript for content, interpretation of clinical and cognitive data. Ms. Huyghe: revising the manuscript for content, laboratory analyses and interpretation of clinical and cognitive data. Dr. Gerard: revising the manuscript for content and data acquisition. Ms. Quenon: revising the manuscript for content, and data acquisition. Dr. Picard: revising the manuscript for content, acquisition of clinical data. Ms. Salman: revising the manuscript for content and data acquisition. MS. Durand: revising the manuscript for content, acquisition of clinical data. Dr. Bedel: revising the manuscript for content, laboratory analysis. Dr. Auriacombe: revising the manuscript for content, acquisition of clinical data. Dr. Lhommel: revising the manuscript for content, interpretation of neuroimaging data. Ms. David: revising the manuscript for content and laboratory analysis. Dr. Kienlen-Campard: revising the manuscript for content, interpretation of data. Dr. Ivanoiu: revising the manuscript for content, interpretation of clinical and cognitive data. Dr. Douxfils: revising the manuscript for content, study concept and design, statistical analysis, interpretation of data. Dr. Levy: revising the manuscript for content, interpretation of clinical data. Dr. Lamari: revising the manuscript for content and laboratory analysis. Dr. Hanseeuw: drafting and revising the manuscript for content, study concept and design, interpretation of data, acquisition of clinical and imaging data, study supervision and coordination, obtaining funding.

