## Supplementary Tables for "Robustness of plasma p-Tau217 diagnostic thresholds for Alzheimer’s disease across various clinical populations and laboratory environments"

| Biomarker | Youden Cutoff | Sensitivity  (CI_95_) | Specificity  (CI_95_) | 95% Se cutoff | 95% Sp cutoff | 95% gray zone (NEG & POS)^1^ | 97.5% Se cutoff | 97.5% Sp cutoff | 97.5% gray zone (NEG & POS)^2^ | AUC ROC |
| --- | --- | --- | --- | --- | --- | --- | --- | --- | --- | --- |
| p-Tau217 (pg/mL) | 0.185 | 94.6%  (IC95: 86.9- 97.9%) | 93.0%  (IC95: 81.4-97.6%) | 0.136 | 0.236 | 12.8%  (7.7 & 5.1%) | 0.128 | 0.283 | 19.7%  (9.4 & 10.3%) | 0.975 |
| AB42/AB40 | 0.0933 | 82.4%  (IC95: 66.5%-91.7%) | 79.2%  (IC95: 59.5-90.8%) | 0.1092 | 0.0724 | 84.5%  (53.4 & 31.1%) | 0.1155 | 0.0724 | 87.9%  (53.4 & 34.5%) | 0.798 |
| p-Tau181 (pg/mL) | 1.920 | 71.9%  (IC95: 54.6-84.4%) | 78.3%  (IC95: 58.1-90.3%) | 1.200 | 3.150 | 74.5%  (38.2 & 36.4%) | 1.100 | 5.670 | 89.1%  (40.0 & 49.1%) | 0.783 |
| p-Tau217/  AB42 | 0.006298 | 94.1%  (IC95: 80.9-99.0%) | 91.7%  (IC95: 74.2-98.5%) | 0.004742 | 0.012568 | 27.6%  (6.9 & 20.8%) | 0.004742 | 0.01525 | 31.0%  (6.9 & 24.1%) | 0.962 |
| p-Tau217/  AB42/  AB40 | 2.096 | 94.1%  (IC95: 80.9-99.0%) | 91.7%  (IC95: 74.2-98.5%) | 1.436 | 3.735 | 24.1%  (5.2 & 19.0%) | 1.323 | 3.840 | 31.0%  (10.3 & 20.7%) | 0.963 |

**Supplementary Table 1: Diagnostic performance of BBMs for predicting clinical-biological AD diagnosis in the Paris validation clinical cohort.**

Additional thresholds corresponding to 95% and 97.5% sensitivity (Se) and specificity (Sp) are also shown. The percentage of individuals falling into the "gray zone" between the 95% Se and 95% Sp thresholds is indicated (^1^), as well as between the 97.5% Se and 97.5% Sp thresholds (^2^). NEG = proportion of subjects between 95%Se cutoff and the Youden cutoff (^1^) or between the 97.5%Se cutoff and the Youden cutoff (^2^). POS = proportion of subjects between the Youden cutoff and 95%Sp cutoff (^1^) or between the Youden cutoff and 97.5%Sp cutoff (^2^).

**A) Primary diagnosis (mutually exclusive)**

| Primary diagnosis | n (%) |
| --- | --- |
| Alzheimer’s disease (probable or highly probable) | **66 (56.4)** |
| Highly probable AD | 53 (45.3) |
| Probable AD | 13 (11.1) |
| Other neurodegenerative disorders | **29 (24.8)** |
| LBD | 7 (6.0) |
| amnestic FTLD | 4 (3.4) |
| iNPH | 4 (3.4) |
| Undetermined non-AD NDD | 4 (3.4) |
| PPA-M/U | 2 (1.7) |
| bvFTD | 2 (1.7) |
| PSP | 2 (1.7) |
| 4R syndrome | 1 (0.9) |
| sbvFTD | 1 (0.9) |
| nfvPPA | 1 (0.9) |
| PD-MCI | 1 (0.9) |
| Non-neurodegenerative / mixed / other etiologies | **22 (18.8)** |
| Anxiety disorder | 6 (5.1) |
| Epileptic disorder | 3 (2.6) |
| Vascular Cognitive Disorders | 3 (2.6) |
| OSA | 2 (1.7) |
| MS | 1 (0.9) |
| Fibromyalgia | 1 (0.9) |
| Undetermined Cognitive Disorder | 1 (0.9) |
| Adrenoleukodystrophy (ABCD1 gene) | 1 (0.9) |
| Schizophrenia | 1 (0.9) |
| CAA | 1 (0.9) |
| Radiation-induced leukoencephalopathy | 1 (0.9) |
| Functional disorder | 1 (0.9) |

**B) Secondary/comorbid diagnosis (not mutually exclusive)**

| Secondary/comorbid diagnosis | n (%) |
| --- | --- |
| Any secondary/comorbid diagnosis | **24 (20.5)** |
| AD | 8 (6.8) |
| LBD | 6 (5.1) |
| CAA | 3 (2.6) |
| LATE | 3 (2.6) |
| Vascular Cognitive Disorders | 2 (1.7) |
| FTLD | 2 (1.7) |
| Undetermined non-AD NDD | 1 (0.9) |

Values are n (% of cohort). Primary diagnoses are mutually exclusive. Secondary/comorbid diagnoses are not mutually exclusive. Abbreviations: AD = Alzheimer’s disease; LBD = Lewy body disease; FTLD = frontotemporal lobar degeneration; PSP = progressive supranuclear palsy; PD = Parkinson’s Disease; PPA-M/U = mixed and/or undetermined primary progressive aphasia; nfvPPA = non-fluent variant primary progressive aphasia; bvFTD = behavioural variant frontotemporal dementia; sbvFTD = semantic behavioural variant frontotemporal dementia; iNPH = idiopathic normal pressure hydrocephalus; CAA = cerebral amyloid angiopathy; LATE = limbic-predominant age-related TDP-43 encephalopathy; MS = Multiple Sclerosis; NDD = Neurodegenerative Disorders; OSA = obstructive sleep apnea.

**Supplementary Table 2.** **Diagnostic distribution in the Paris routine-care cohort (n=117)**

| Plasma  CSF | Negative | 97.5% gray zone | Positive | Total |
| --- | --- | --- | --- | --- |
| A+T+ | 0 | 13 | 48 | **61** |
| A+T- | 0 | 4 | 9 | **13** |
| A-T+ | 3 | 2 | 0 | **5** |
| A-T- | 27 | 9 | 2 | **38** |
| Total | **30** | **28** | **59** | **117** |

**Supplementary Table 3**: **Usefulness of CSF as a 2^nd^ line AD biomarker evaluation in clinical practice.** Contingency table showing plasma and CSF status in the Paris validation clinical cohort, using double plasma ptau217 cutoffs derived from the Brussels Exploration cohort with 97.5% sensitivity and specificity.

| Plasma  CSF | Negative | 95% gray zone | Positive | Total |
| --- | --- | --- | --- | --- |
| A+T+ | 3 | 7 | 51 | **61** |
| A+T- | 1 | 2 | 10 | **13** |
| A-T+ | 3 | 2 | 0 | **5** |
| A-T- | 29 | 6 | 3 | **38** |
| Total | **36** | **17** | **64** | **117** |

**Supplementary Table 4**: **Usefulness of CSF as a 2^nd^ line AD biomarker evaluation in clinical practice.** Contingency table showing plasma and CSF status in the Paris validation clinical cohort, using double plasma ptau217 cutoffs derived from the Brussels Exploration cohort with 95% sensitivity and specificity.
